# Patterns of Racial and Socioeconomic Inclusivity of Hospital Care Among the Medicare Population

**DOI:** 10.1101/2021.05.24.21257551

**Authors:** Vikas Saini, Kelsey Chalmers

**Author notes:** Corresponding Author: Vikas Saini, M.D., Lown Institute, 163 Highland Ave, Needham, MA 02494.

## Abstract

**Importance:** Measures of health care disparities across racial and socioeconomic groups are needed to guide research and policy, particularly for hospitals, because they are the major locus of health care.

**Objective:** To quantify socioeconomic differences between a hospital’s patients and the surrounding area, and to assess associations with hospital characteristics.

**Design:** A cross-sectional observational study using Medicare fee-for-service claims and the 2019 American Community Survey (ACS) 5-year zip code data. Quantile regressions identified hospital characteristics associated with lowest and highest inclusivity.

**Setting:** Inpatient admissions to non-specialty, non-federal hospitals (N = 3,426) in calendar years 2018 and 2019.

**Participants:** Medicare beneficiaries enrolled in Parts A & B, and over the age of 65 years

**Main Outcome and Measures:** Inclusivity was defined as the differences between education level, income, and proportion of racial groups, between a hospital’s community area and its patient zip codes (obtained from US Census data). The community area was calculated based on a hospital’s patient counts from contributing zip codes and by finding this travel time radius from each hospital campus. Percentiles of the inclusivity scores and their composite were compared across hospital characteristics using quantile regressions.

**Results:** We included 10,221,387 patients with a mean (± standard deviation) age of 77.4 (8.0) years and 55.3% were female. The median travel time radius was 48.2 minutes (IQR: 34.7 to 75.0). Weighted median incomes for hospitalized patients’ zip codes ranged from $27,060 less than their community area to $31,505 more. Black people had the widest weighted percentage differences across hospitals: one hospital had a Black patient proportion 41.6% greater than its community area, while another had a proportion 37.2% less than its community area. Safety net status was the characteristic consistently associated with a higher median inclusivity score (0.4 points greater than non-safety net hospitals, CI: 0.31 to 0.45; p < 0.001). Metropolitan area hospitals had both higher 90^th^ and lower 10^th^ percentile scores compared to non-metropolitan, as did teaching hospitals compared to non-teaching.

**Conclusions and Relevance:** US hospitals’ measured inclusivity varies widely, with patterns of greater segregation in metropolitan areas. The results for safety net hospitals reflect their essential role in the US healthcare system.

**Key points:** *Question:* What are the variation patterns of socioeconomic and racial characteristics of hospital inpatient populations in relation to the demographics of their surrounding communities?

*Findings:* Using Medicare claims data, we calculated the differences in race, income and educational attainment between hospitalized patients and a defined surrounding community area. Quantile regressions showed that the most and the least inclusivity existed in larger, teaching, and metropolitan area hospitals. Safety net hospitals consistently maintained higher inclusivity across both urban and rural settings.

*Meaning:* Systematic patterns of racial and class segregation exist among hospitalized Medicare patients.

## Introduction

Various methods describe hospital ‘market areas’ and examine hospital care through a lens of market competition.^1,2^ To our knowledge, such methods have not been applied to examine socioeconomic and racial disparities in hospital use across geographic areas. Hospitals account for one third of US health care spending, representing a critical locus for assessing disparities^3–6^ which can have impact on a broad range of health outcomes.^7–9^

Metrics of structural racism and income inequality are associated with disparities in clinical outcomes but to our knowledge, none report at the hospital level.^10-11^ The Index of Dissimilarity, originally applied to the measurement of residential segregation^12,13^ can be applied to hospital segregation within geographic regions,^14^ but not for individual hospitals. We sought to develop such a hospital-level metric, hospital *inclusivity*, using national administrative claims data and an empirically derived definition of market area (the hospital *community area*).

## Methods

### Data sources

We used a 100% sample of fee-for-service claims from the Centers for Medicare & Medicaid Services’ (CMS). We counted unique inpatient visits for beneficiaries 65 years and older by zip code from the Medicare Provider Analysis and Review (MedPAR) table at short-term general and critical access hospitals in 2018 to 2019. We excluded hospitals that were not on the 2020 CMS Hospital Compare website,^11^ federal, specialty, or had fewer than 50 inpatient visits in this period. Hospital address coordinates were from HealthData.gov.^15^

Zip codes were mapped to zip code tabulation area (ZCTA), and excluded patients without a ZCTA mapping. We used the 2019 American Community Survey (ACS) 5-year variables.^16^ This included the ZCTA median household income in the past 12 months (in 2018 inflation-adjusted dollars) where the head of household was 65 years and over; the proportion of persons aged 65 and over in the ZCTA with an education level less than 9th grade, 9th to 12th grade, high school diploma, some college (no degree), associate’s degree, bachelors’ degree, graduate or professional degree; and the proportion of persons aged 65 and over that were American Indian or Alaska Native, Asian, Black or African American, Hispanic or Latino, Native Hawaiian and other Pacific Islander, Other, Two or more races, and White (not Hispanic or Latino). We calculated a continuous ZCTA education value by assigning each level a value from from 1 to 7 and finding the median.

We replaced 99.7% of 3,508 ZCTAs with missing median incomes with the county-level equivalent, and for the remaining we used predictive mean matching to impute based on the state, the percentage of people in the state over 65, and the percentage over 65 with a bachelors’ degree.

We used *OpenStreeMap*^17^ data to estimate travel time to ZCTA centroids from hospital locations.

### Community area

We first found the furthest ZCTA from which a large enough proportion of a hospital’s patients lived (for algorithm details see online Supplementary Appendix), using the geodesic distance from the campus address to ZCTA centroids.^18^ For hospitals with multiple campuses, we matched ZCTAs with patient counts to the nearest campus and then found the furthest ZCTA for each campus.

We then found the estimated driving travel time using *OpenStreetMaps* data and the *osrm* R package^19^ between each campus and its furthest ZCTA. We defined the hospital community area (CA) as the isochrone (the area with a boundary of equal travel time) from each campus with this travel time. ZCTAs overlapping isochrone boundaries were partially included in the CA demographics as a fraction of their total area included in the isochrone.

### Community area and hospital patient demographics

ZCTAs demographics within the CA were weighted based on the geodesic distances to a hospital. ZCTAs within a radius including at least 50% of the hospital’s patients (the 50^th^ percentile radius) were equally weighted. Otherwise, ZCTAs were decreasingly weighted the further they were from the hospital, based on the decay rate which gave a weight value at the furthest ZCTA from the hospital proportional to the number of patients outside the CA. For hospitals with multiple campuses, we defined the hospital ‘location’ as the weighted average coordinates based on the number of patients in each ZCTA and used this to calculate the distance from each ZCTA location and its resulting CA weight.

Equation 1 shows the estimate of the CA income, *CI*_*H*_ for hospital *H*, across *N* zip codes in the CA, each with weighting *w*_*i*_, population over 65 *P*_*i*_ and median income *inc*_*i*_.

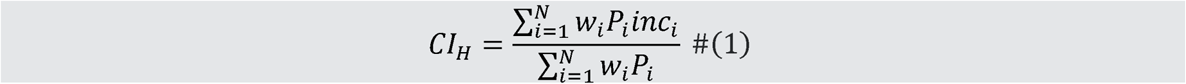

We used the same approach for the CA education estimate.

For the CA racial estimates (equation 2), we found the weighted proportion of persons in each group (for *k*= 1,…,8) across CA ZCTAs.

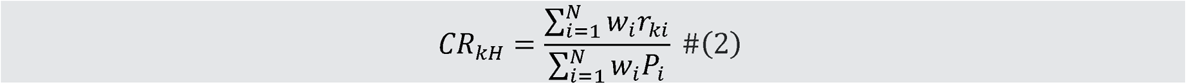

Where *r*_*ki*_ is the number of persons 65 and over in group *k* in ZCTA *i*.

The hospital income (*I*_*H*_), education (E_*H*_) and racial (*R*_*kH*_) estimates were the demographics of CA ZCTAs, weighted by the hospital’s patient counts from each ZCTA.

### Inclusivity metrics

The education and income inclusivity metrics were the log-ratio of the CA to hospital estimates. We used a similar measure to Chi-squared difference for racial inclusivity (equation 3). We did not include the proportion of white people in equation 3 in order to give hospitals a higher inclusivity score if they had a higher estimated patient proportion of racialized groups relative to their CA.

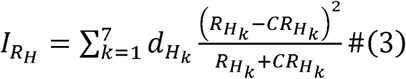

Where 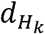 is given by:

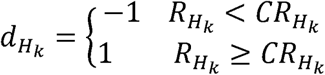

### Composite inclusivity score

The composite inclusivity score was the sum of normalized racial, income and education inclusivity scores. Racial inclusivity scores were not included in the composite for hospitals with a homogeneous CA, defined as areas where probability that two randomly selected persons being the same race was greater than 95%. Scores were normalized by min-max transformation.

### Hospital characteristics

Safety net hospitals were the top 20% of hospitals by proportion of inpatient days billed as dual eligible. Other hospital characteristics were defined using the American Hospital Association 2020 dataset, and included size (based on number of beds), teaching status, core-based statistical area (CBSA) region type, and profit status. Major teaching hospitals are listed as a Member of Council of Teaching Hospitals of the Association of American Medical Colleges; minor teaching hospitals lacked these memberships, but had a medical school affiliation. CBSA types were defined as either metropolitan (metro) with 50,000 or more people, micropolitan (micro) with 10,000 to 50,000 people, or rural areas. Government, nonfederal and nongovernment, not-for-profit were labeled as non-profit hospitals.

### Statistical analysis

We report descriptive statistics of the community area travel radius, the distance of the furthest ZCTA included in the community area, the 50^th^ percentile radius, and the inclusivity scores. We used quantile regressions to investigate differences across hospital characteristics.^20,21^ We inspected the range of inclusivity scores within each Hospital Referral Regions (HRR)^22^ and identified the HRRs where safety net status had the greatest impact on the mean inclusivity score using multi-level regression, with HRR as a random effect.^23^

The analysis took place between January to March 2022 using SAS Enterprise Guide version 7.15 HF8 and R version 4.0.5 (with data manipulation using *tidyverse* package^24^ and plotting using *ggplot*^25^).

This study was deemed exempt from IRB review and a waiver of consent was granted as there was no risk to patients.

## Results

Appendix Table 1 shows the characteristics of the 3,426 hospitals in our study, with 10,221,387 included patients. The mean age of patients was 77.4 (standard deviation [SD] of 8.0), with 8,128,568 white patients (79.5%) and 5,652,950 women (55.3%).

The median hospital CA travel radius was 48.2 minutes (IQR: 34.7 to 75.0). The median distance between hospitals and their furthest CA ZCTA was 37.3 miles (IQR: 26.3 to 60.2). Micro-area hospitals had the highest median travel radius (51.5 minutes [IQR: 38.8 to 75.9]) compared to metro- and rural hospitals. Extra large hospitals had the greatest travel radius (median 76.0 minutes [IQR: 40.7 to 120.8]), with extra small hospitals having the smallest (median 41.2 minutes [IQR: 32.4 to 55.7]). Rural hospitals had the lowest 50th patient percentile radius (median 5.5 miles [IQR: 2.2 to 9.6]) compared to other hospitals. The mean proportion of patients within hospital CAs was 91.2% [SD 5.8] (median 92.4%, IQR: 89.4 to 94.6).

### Inclusivity scores

The median composite inclusivity score was -0.03 points (IQR: [-0.37, 0.44)]). The median score for income inclusivity was -0.07 points (IQR [-0.44, 0.46]), education was -0.07 points (IQR [-0.44, 0.47]), and racial inclusivity was 0.23 points (IQR [-0.21, 0.33]). Figure 1 shows the differences between the CA and hospitals’ patient demographics. The maximum positive difference in the income estimate, reflecting greater inclusivity, was $31,505 and the largest negative difference was -$27,060. For education, the largest differences between the median education levels were 1.8 and -1.1, approximately equivalent to two adjacent education levels. Among racialized groups, Native Hawaiian or Pacific Islander populations had the smallest differences ranging from 5.0% greater to 0.6% less than the CA. The largest differences were seen for Black populations, where one hospital had a maximum difference of 41.6% greater than its CA while another had 37.2% less than its CA (see Appendix Table 2 for details).

**Figure 1.**
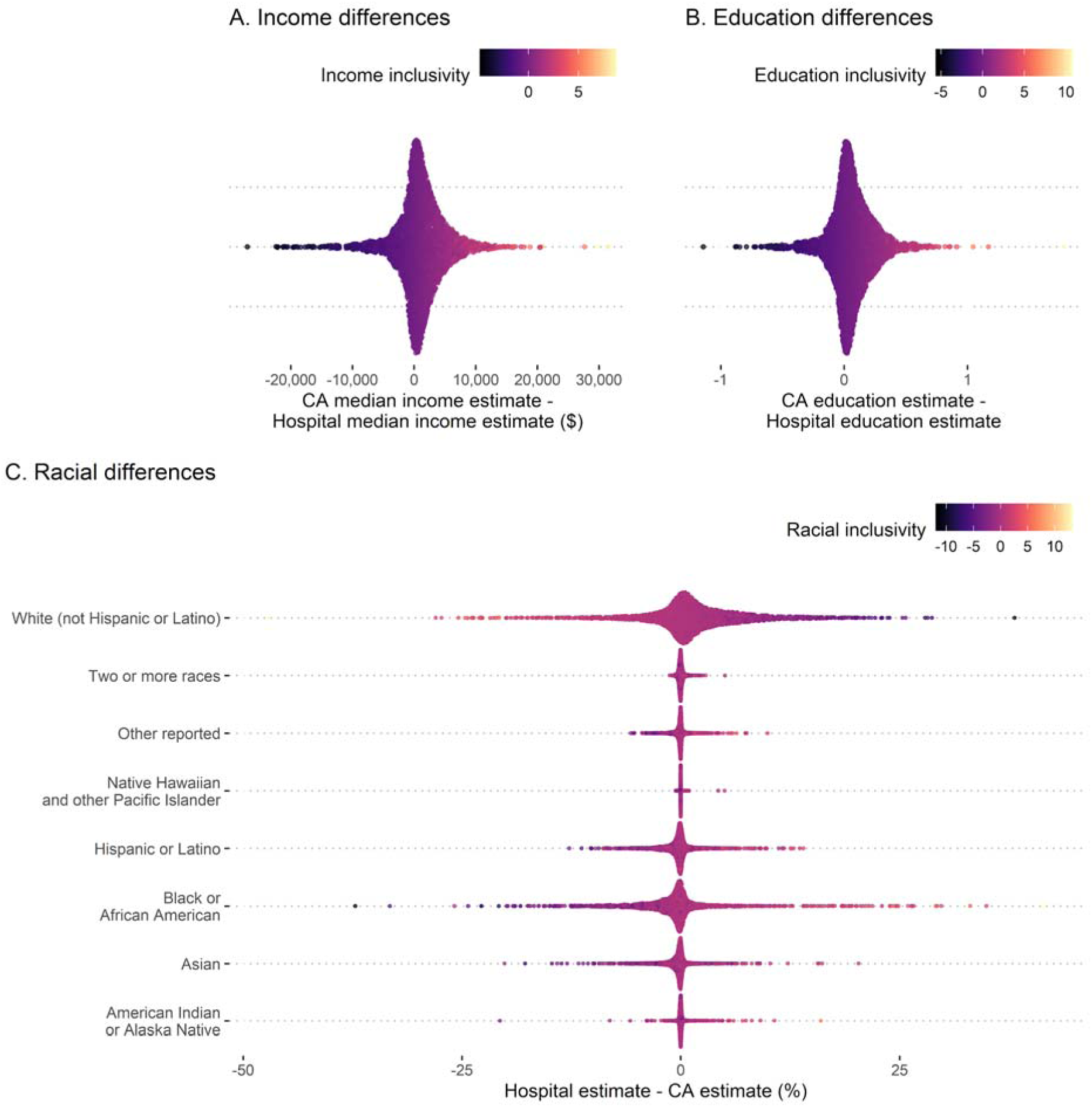
Individual hospital results of the differences between the hospital demographic estimates and the community area (CA) estimates. A. median income, B. educational attainment and C. percentage of each racial group. Points are shown for every hospital (N = 3,548), with the relative density of these points shown on the y-axis. Hospital points are colored based on each inclusivity score metric.

Inclusivity scores were significantly and positively correlated with each other within metro-area hospitals (Figure 2). Income and education inclusivity correlation for metro-area hospitals (rank correlation R = 0.61; p < 0.001) was higher than each with racial inclusivity (R = 0.20; p < 0.001 income; R = 0.16; p < 0.001 education). Racial inclusivity was not correlated with either income or education in rural hospitals, but income and education were (R = 0.39; p < 0.001).

**Figure 2.**
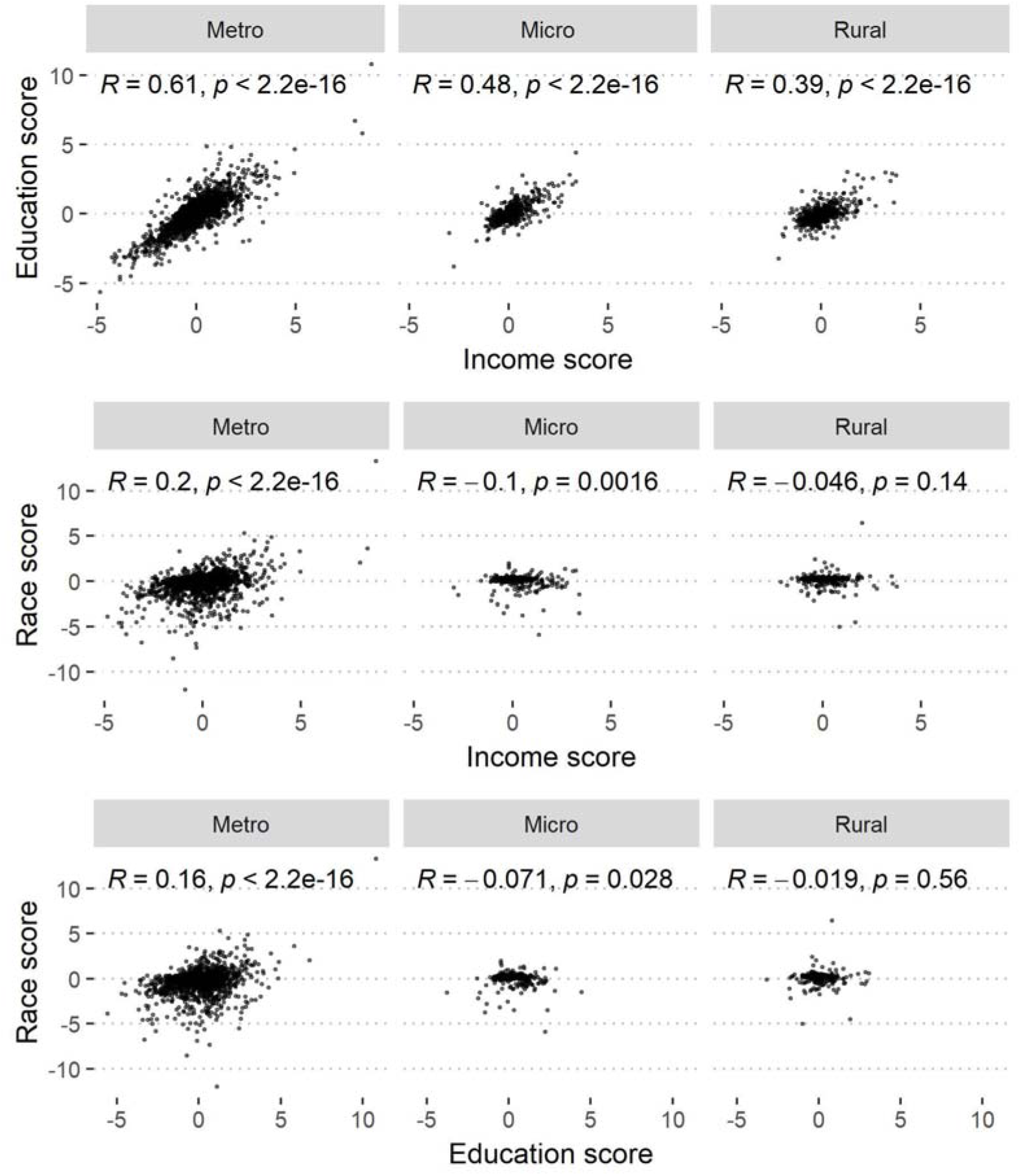
Comparison of the three inclusivity scores across hospitals: income, education and race. The Kendall rank correlation coefficient and p-value for the statistical test that this correlation is not zero between each variable pair is shown.

### Inclusivity association with hospital characteristics

Figure 3 shows the unadjusted 10^th^ to 90^th^ inclusivity score percentiles by hospital characteristics. Metropolitan, major teaching and large hospitals had the largest range of inclusivity. The estimated composite score quantile regression coefficients are shown in Appendix Figure 1, with the adjusted 10^th^, 50^th^ and 90^th^ percentile estimates reported in Appendix Table 3.

**Figure 3.**
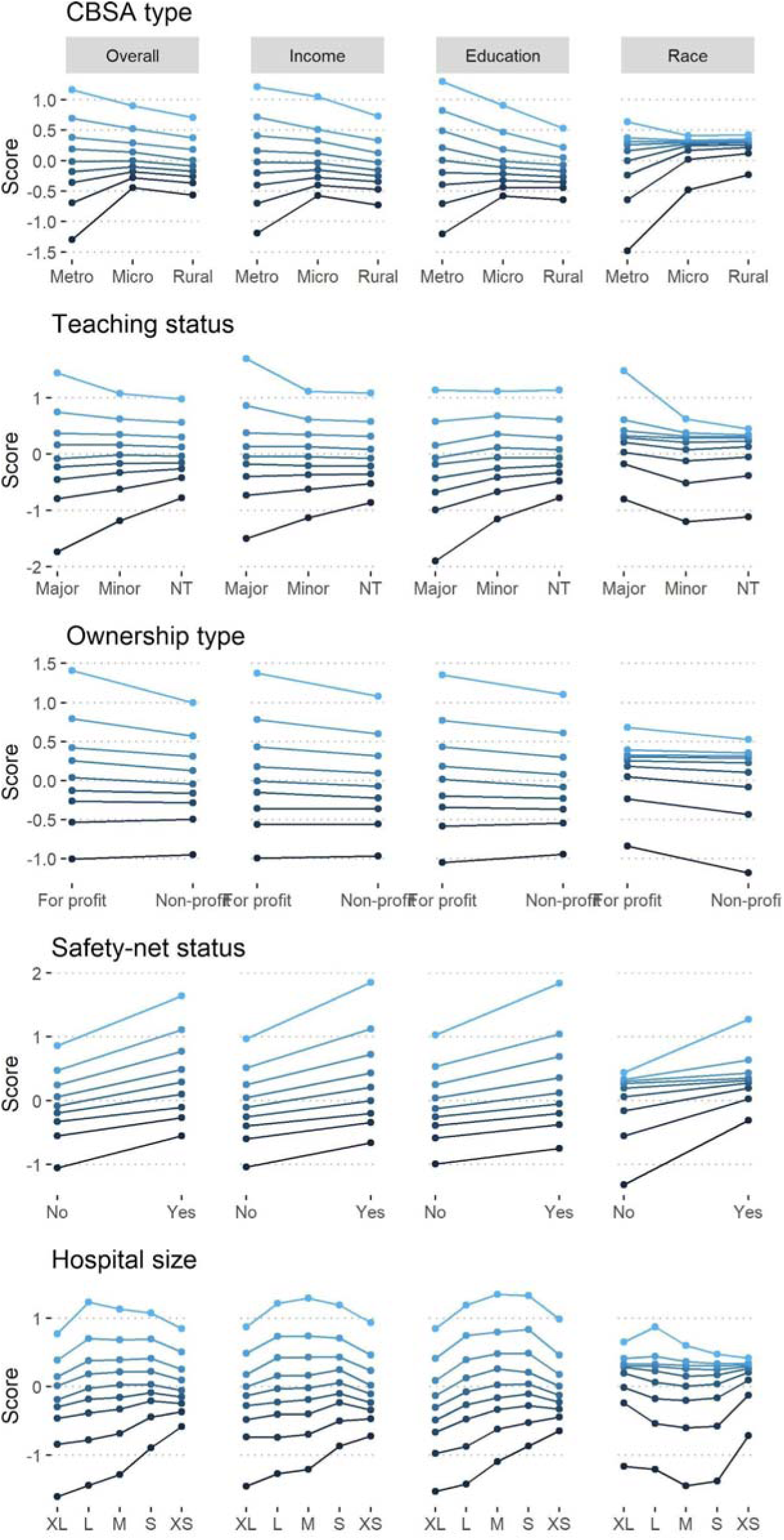
The overall, income, education and race inclusivity scores by hospital characteristics. Points show the 10^th^ to 90^th^ percentile scores for each hospital characteristic level, with lines drawn between the respective percentiles for each hospital characteristic level (with each line a different color shaded from lowest quantile to highest). NT: non-teaching; FP: for profit, NP: non-profit; XL: extra large; L: large; M: medium; S: small; XS: extra small.

The 90^th^ percentile of composite inclusivity of metropolitan, major teaching, and safety net hospitals were greater than the 90^th^ percentile inclusivity of other hospital types. Metropolitan hospitals had an adjusted 90^th^ percentile score 0.46 points greater than rural hospitals (CI: 0.25 to 0.66; p < 0.001). The 10^th^ percentile scores of metropolitan, major teaching and non-safety net hospitals were lower than the 10^th^ percentile scores of other hospital types. Major teaching hospitals were associated with a greater composite score range compared to minor teaching and non-teaching hospitals, with no significant differences across median scores (Appendix Figure 1), as was metropolitan compared to non-metropolitan hospitals. Safety net hospital percentile estimates were greater than non-safety net hospitals across all quantile regressions.

The online supplement also shows the quantile regressions for income, education, and the racial inclusivity scores (Appendix Figure 2).

### Inclusivity by hospital referral regions

The difference between the maximum and minimum inclusivity within an HRR ranged from 0.02 points (Lynchburg, Virgina) to 15.2 points (Fort Lauderdale, Florida). Figure 4 shows the map of these differences, with the largest visible in metropolitan areas.

**Figure 4.**
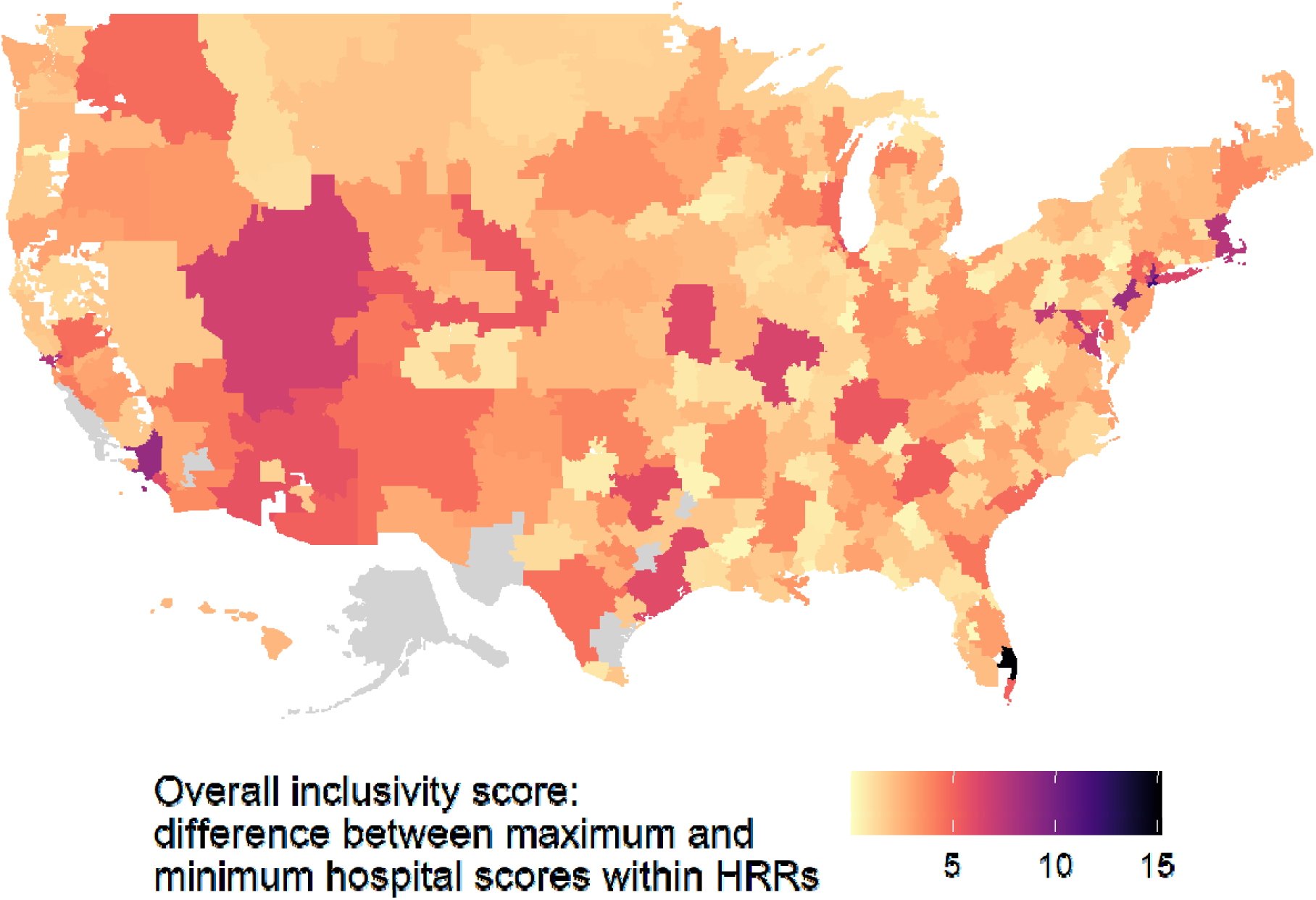
The range of composite inclusivity scores across hospitals within each HRR (The Maximum - Minimum Inclusivity Scores), showing the degree in dispersion of inclusivity and patterns of local segregation in hospital utilization among Medicare beneficiaries within certain regions of the country.

Twenty-four HRRs had a significant difference between the average inclusivity scores of safety net and non-safety net hospitals, mostly in metropolitan areas (Appendix Figure 3). Within rural areas, safety net hospitals had significantly higher median inclusivity scores for income and education (Appendix Table 4).

## Discussion

Due to persistent injustices of racism and economic inequality, researchers, policymakers and communities have called for metrics to promote accountability and ensure lasting system change.^26^ With this novel metric we measure hospitals’ service to their surrounding communities, and define inclusivity as the deviance between the hospital patient demographics to its CA demographics. Hospitals were deemed more *inclusive* if they had a higher patient population from areas with lower income, lower education, or more racialized groups compared to the CA population. By design, all hospitals that had a patient population which exactly matched their CA would have a ‘neutral’ inclusivity score of zero. The median education and income inclusivity scores were negative while the racial inclusivity scores were positive, which may reflect different hospitalization rates by socioeconomic status.^27,28^

We calculated inclusivity metrics of education, income and race, as the intersections of these may be important in understanding exclusion patterns. We based racial inclusivity on the sum of percentage differences for each racialized group reported in the ACS. Most hospitals with a low racial inclusivity score had higher admissions from areas with more white people relative to their CA. Some hospitals, however, had a low racial inclusivity score if negative differences across one or more racialized groups was greater than the positive differences across other racialized groups. We did not combine all racialized groups into one because we decided to keep this level of granularity. The low correlations between racial and income or education inclusivity, relative to income versus education inclusivity, may partly arise from this decision. Hospitals with high proportions of one racialized group but not others within its CA would have lower racial scores, potentially unrelated to their patients’ income.

Hospitals’ inclusivity scores wide range provides insight into the local distribution of racial and socioeconomic differences amongst Medicare beneficiaries. While the median inclusivity scores did not vary substantially by hospital characteristics (other than safety net status), the quantile regressions showed that extreme inclusivity scores were associated with particular hospital types. The range of hospital inclusivity within HRRs also varied nationally.

Our finding that the range of inclusivity scores is greatest in metropolitan hospitals is not surprising. These inclusivity differences likely reflect diversity in metro-areas. This diversity, however, does not necessarily lead to differences in this inclusivity score; that it does likely reflects how socioeconomic factors, both current and historical, determine health care utilization. These include residential segregation, itself coupled to historical sequences of immigration, labor mobility, and the linkages of insurance coverage to differing employment conditions and partitions in the labor and health insurance markets. Inclusivity as reported here may enable further investigation of these additional associations.

We defined safety net hospitals as those with the highest proportion of dual eligible patient days, and found that these hospitals were associated with higher inclusivity scores compared to non-safety net hospitals. This finding is not tautological because a safety net hospital could in theory have a low or even negative inclusivity score. The positive inclusivity scores for safety net hospitals demonstrate that they disproportionately and selectively serve the lower-income ZCTAs within larger areas containing both high- and low-income neighborhoods. Safety net hospitals are likely complementary to nearby hospitals with lower inclusivity scores selectively serving high-income neighborhoods. While this may provide access to hospital care for the entire community population, it is unlikely to be an adequate or just distribution of health care resources.

The inclusivity scores are dependent on the defined CA. Instead of a fixed radius method,^1^ we used a method similar to the variable radius methods of previous analyses of hospital market areas.^29^ Ours differed in that it was defined by an algorithm sensitive to the distribution characteristics of each hospital rather than by a single cut-off for all hospitals at a targeted proportion of patients. Nevertheless, our CA encompassed on average 91.2% of a hospital’s patients (median 92.4%) similar to the defined radius used in previous work.^2^

Since we used Medicare data, our results do not reflect patient choice based on insurance coverage, nor would they reflect hospital marketing towards patients with more lucrative commercial insurance coverage. The pattern of disparities in inclusivity we report are more likely reflections of patients and hospital habits and attitudes, driven by such issues as trust,^30^ the communities a hospital seeks to engage, patterns of patient preference and utilization dating to pre-Medicare insurance status,^31^ rates of supplemental insurance,^32^ hospital financial aid and collection policies, and factors such as the number of Black or minority doctors.^33^ Policy makers will have to address these challenges if they wish to improve the inclusivity of care among the hospitalized Medicare population.

## Limitations

This study included Medicare fee-for-service patients only, and may not apply to other populations such as Medicare Advantage, commercially insured, or uninsured patients.

We used CBSA definitions of population areas. Our results, particularly in comparing the CA sizes, might have been different had we used finer gradations of population density.

Our calculations assumed that everyone in a particular ZCTA acts with the same propensity to use a hospital. This does not consider socioeconomic and cultural changes within ZCTAs, such as gentrification of poorer neighborhoods.

Another limitation is that we did not use demographic data at the patient level, but at the ZCTA level. While patient-level data on race exists in the Medicare files, we decided to use ZCTA to be consistent with the income and education values, which were only available at a ZCTA level.

## Conclusions

We present a method of quantifying socioeconomic and racial inclusion in US hospitals. Inclusivity, the degree to which a hospital’s patients’ racial and socioeconomic characteristics reflect its community area, varied widely. Inclusivity varied most in metropolitan and major teaching hospitals. Safety net hospitals’ inclusivity scores show they serve an important social function in both urban and rural settings. These inclusivity metrics hold promise for identifying associations with other factors, but more research is required. They represent an objective, transparent method of measuring one specific aspect of inequities in health care. Inclusivity assessed over time may provide a tool for the measurement of the effects of policy actions designed to mitigate structural racism and socio-economic disparities in US hospital care.

## Data Availability

Medicare data was accessed after approvals and agreement with CMS. Individual hospital results will be available on https://lownhospitalsindex.org/. Other data can be accessed upon reasonable request to The Lown Institute, please contact.

