## Appendix for "Patterns of Racial and Socioeconomic Inclusivity of Hospital Care Among the Medicare Population"

### Appendix Table 1. The characteristics of hospitals, patients, and Community Areas in our sample.

|  | **CBSA of hospital** | | | |
| --- | --- | --- | --- | --- |
|  | All (N = 3,426) | Metro (N = 2,149) | Micro (N = 558) | Rural (N = 719) |
| **Hospitals** (N, %) |  |  |  |  |
| Ownership type |  |  |  |  |
| For profit | 469 (14) | 361 (17) | 64 (11) | 44 (6.1) |
| Non-profit | 2,957 (86) | 1,788 (83) | 494 (89) | 675 (94) |
| Teaching class |  |  |  |  |
| Major | 218 (6.4) | 214 (10.0) | 4 (0.7) | 0 (0) |
| Minor | 1,123 (33) | 941 (44) | 126 (23) | 56 (7.8) |
| Non-teaching | 2,085 (61) | 994 (46) | 428 (77) | 663 (92) |
| Hospital type |  |  |  |  |
| Acute Care | 2,514 (73) | 1,960 (91) | 387 (69) | 167 (23) |
| Critical Access | 912 (27) | 189 (8.8) | 171 (31) | 552 (77) |
| Safety net status |  |  |  |  |
| Non-safety net | 2,778 (81) | 1,787 (83) | 473 (85) | 518 (72) |
| Safety net | 648 (19) | 362 (17) | 85 (15) | 201 (28) |
| Bed size |  |  |  |  |
| Extra small | 1,368 (40) | 357 (17) | 327 (59) | 684 (95) |
| Small | 461 (13) | 297 (14) | 133 (24) | 31 (4.3) |
| Medium | 733 (21) | 655 (30) | 74 (13) | 4 (0.6) |
| Large | 631 (18) | 608 (28) | 23 (4.1) | 0 (0) |
| Extra large | 233 (6.8) | 232 (11) | 1 (0.2) | 0 (0) |
| **Patients** (N) | 10,221,387 | 9,433,117 | 783,309 | 284,647 |
| Age (mean, SD) | 77.4 (8.0) | 77.3 (8.0) | 77.7 (8.0) | 78.7 (8.2) |
| Sex, Female (N, %) | 5,652,950 (55.3) | 5,196,477 (55.1) | 441,403 (56.4) | 165,811 (58.3) |
| Race* (N, %) |  |  |  |  |
| Non-Hispanic white | 8,128,568 (79.5) | 7,410,305 (78.6) | 711,840 (90.9) | 260,340 (91.5) |
| Black or African-American | 987,964 (9.7) | 946,685 (10.0) | 41,768 (5.3) | 14,954 (5.3) |
| Asian/Pacific Islander | 219,747 (2.1) | 216,577 (2.3) | 3,404 (0.3) | 469 (0.2) |
| Hispanic | 664,265 (6.5) | 653,752 (6.9) | 10,936 (1.4) | 3,765 (1.3) |
| American Indian or Alaska Native | 35,537 (0.3) | 29,657 (0.31) | 5,880 (0.8) | 2,483 (0.9) |
| Other | 68,903 (0.7) | 65,538 (0.7) | 3,495 (0.4) | 919 (0.3) |
| Unknown | 116,403 (1.1) | 110,603 (1.2) | 5,986 (0.8) | 1,717 (0.6) |
| **CA travel time radius** (minutes) |  |  |  |  |
| Median (IQR) | 48.2 (34.7, 75.0) | 49.8 (34.1, 81.2) | 51.5 (38.8, 75.9) | 42.0 (34.0, 58.4) |
| Mean (SD) | 61.9 (45.0) | 65.2 (50.1) | 61.6 (33.8) | 50.8 (29.2) |
| Range | 2.6, 526.9 | 2.6, 526.9 | 12.4, 238.9 | 4.8, 272.8 |
| Comparison (p-value) | <0.001 |  | | |
| **Distance of furthest ZCTA included in CA** (miles) |  |  |  |  |
| Median (IQR) | 37.3 (26.3, 60.2) | 38.7 (25.9, 65.7) | 40.9 (29.2, 59.7) | 33.1 (25.6, 45.4) |
| Mean (SD) | 49.7 (39.4) | 53.5 (45.2) | 48.4 (27.9) | 39.4 (22.8) |
| Range | 0.9, 474.3 | 0.9, 474.3 | 11.0, 226.0 | 2.5, 240.7 |
| Comparison (p-value) | <0.001 |  | | |
| **50th percentile radius** (miles) |  |  |  |  |
| Median (IQR) | 6.8 (3.8, 10.3) | 7.0 (4.5, 10.4) | 7.0 (2.7, 10.9) | 5.5 (2.2, 9.6) |
| Mean (SD) | 8.1 (6.7) | 8.7 (7.0) | 7.9 (6.3) | 6.7 (5.8) |
| Range | 0.1, 71.6 | 0.1, 71.6 | 0.1, 45.5 | 0.1, 69.9 |
| Comparison (p-value) | <0.001 |  | | |
| **Patients within CA-R** (%) |  |  |  |  |
| Median (IQR) | 92.4 (89.4, 94.6) | 92.0 (88.7, 94.4) | 93.6 (91.7, 95.2) | 92.5 (90.0, 94.3) |
| Mean (SD) | 91.2 (5.8) | 90.7 (6.3) | 92.8 (3.8) | 91.6 (5.4) |

CBSA: Core based statistical area. CA: community area. ZCTA: zip code tabulation area. *Race recorded in MBSF file using Research Triangle Institute Race Code.

### Appendix Table 2. Community area and hospital estimates of education levels, median income, and percentage of different racial groups based on data from ZCTA-level American Community Survey data.

| Value | **Community area scores** | | | **Hospital scores** | | |
| --- | --- | --- | --- | --- | --- | --- |
|  | **Median (IQR)** | **Mean (SD)** | **Range** | **Median (IQR)** | **Mean (SD)** | **Range** |
| Education level | 3.9 (3.7, 4.1) | 3.9 (0.4) | 1.9, 5.4 | 3.8 (3.6, 4.1) | 3.9 (0.4) | 1.8, 5.5 |
| Median income ($) | 44,074 (38,755, 50,464) | 45,151 (9,418) | 21,990, 99,845 | 42,453 (37,337, 49,217) | 44,205 (10,407) | 18,950, 106,954 |
| American Indian persons (%) | 0.3 (0.1, 0.5) | 0.6 (1.8) | 0.0, 62.4 | 0.2 (0.1, 0.5) | 0.6 (2.2) | 0.0, 90.3 |
| Asian persons (%) | 1.1 (0.4, 2.4) | 2.7 (5.1) | 0.0, 68.5 | 0.8 (0.3, 2.0) | 2.3 (5.0) | 0.0, 68.2 |
| Black or African American persons (%) | 3.9 (0.8, 11.3) | 8.1 (10.6) | 0.0, 82.1 | 2.5 (0.5, 8.8) | 7.4 (11.7) | 0.0, 87.4 |
| Hispanic persons (%) | 1.9 (0.9, 5.9) | 4.9 (7.0) | 0.0, 49.5 | 1.7 (0.8, 5.1) | 4.6 (7.2) | 0.0, 49.5 |
| Other race reported (%) | 0.4 (0.1, 1.4) | 1.1 (1.8) | 0.0, 17.2 | 0.3 (0.1, 1.1) | 1.1 (2.0) | 0.0, 21.9 |
| Native Hawaiian or Pacific Islander persons (%) | 0.0 (0.0, 0.0) | 0.1 (0.3) | 0.0, 9.4 | 0.0 (0.0, 0.0) | 0.1 (0.4) | 0.0, 10.5 |
| Two or more races reported (%) | 0.8 (0.5, 1.1) | 0.9 (0.8) | 0.0, 13.5 | 0.7 (0.5, 1.1) | 0.9 (0.9) | 0.0, 14.4 |
| White, non-Hispanic persons (%) | 86.6 (72.9, 94.8) | 81.6 (16.3) | 13.0, 100.0 | 89.4 (75.3, 95.8) | 83.0 (17.2) | 5.7, 100.0 |

### Appendix Table 3. The unadjusted and adjusted 10^th^, 50^th^ and 90^th^ percentile inclusivity score estimates.

| **Hospital characteristic** | **10^th^ percentile** | | **50^th^ percentile** | | **90^th^ percentile** | |
| --- | --- | --- | --- | --- | --- | --- |
|  | **Unadj.** | **Adjusted (CI)** | **Unadj.** | **Adjusted (CI)** | **Unadj.** | **Adjusted (CI)** |
| **Overall inclusivity score** | | | | | | |
| Hospital size |  |  |  |  |  |  |
| Extra small | -0.6 | -0.7 (-0.9, -0.5) | -0.1 | 0.1 (0.1, 0.2) | 0.8 | 1.5 (1.3, 1.7) |
| Small | -0.9 | -0.7 (-0.9, -0.6) | 0.0 | 0.2 (0.1, 0.3) | 1.1 | 1.4 (1.2, 1.6) |
| Medium | -1.3 | -0.9 (-1.2, -0.6) | 0.0 | 0.2 (0.1, 0.2) | 1.1 | 1.5 (1.3, 1.7) |
| Large | -1.4 | -1.0 (-1.3, -0.8) | 0.0 | 0.1 (0, 0.2) | 1.2 | 1.4 (1.2, 1.7) |
| Extra large | -1.6 | -0.8 (-1.3, -0.4) | -0.2 | -0.1 (-0.1, 0) | 0.8 | 0.9 (0.7, 1.1) |
| CBSA type |  |  |  |  |  |  |
| Metro | -1.3 | -1.2 (-1.4, -1.1) | 0.0 | 0.1 (0.1, 0.2) | 1.2 | 1.5 (1.4, 1.7) |
| Micro | -0.4 | -0.6 (-0.7, -0.4) | 0.0 | 0.1 (0.1, 0.2) | 0.9 | 1.4 (1.2, 1.6) |
| Rural | -0.6 | -0.7 (-0.9, -0.6) | -0.1 | 0.1 (0, 0.1) | 0.7 | 1.1 (0.9, 1.3) |
| Ownership |  |  |  |  |  |  |
| For-profit | -1.0 | -0.8 (-0.9, -0.6) | 0.0 | 0.1 (0, 0.2) | 1.4 | 1.4 (1.2, 1.6) |
| Non-profit | -0.9 | -0.9 (-1, -0.8) | 0.0 | 0.1 (0.1, 0.2) | 1.0 | 1.3 (1.2, 1.5) |
| Teaching class |  |  |  |  |  |  |
| Major | -1.7 | -1.1 (-1.5, -0.8) | -0.1 | 0.2 (0.1, 0.4) | 1.4 | 1.9 (1.5, 2.3) |
| Minor | -1.2 | -0.6 (-0.8, -0.5) | 0.0 | 0.1 (0, 0.1) | 1.1 | 1.1 (0.9, 1.2) |
| Non-teaching | -0.8 | -0.7 (-0.9, -0.6) | 0.0 | 0.1 (0, 0.1) | 1.0 | 1.1 (0.9, 1.2) |
| Safety net status |  |  |  |  |  |  |
| Non-safety net | -1.1 | -1 (-1.1, -0.8) | -0.1 | -0.1 (-0.1, 0) | 0.9 | 1.0 (0.8, 1.1) |
| Safety net | -0.6 | -0.7 (-0.9, -0.6) | 0.3 | 0.3 (0.2, 0.4) | 1.6 | 1.7 (1.5, 1.9) |
| **Income inclusivity score** | | | | | | |
| Hospital size |  |  |  |  |  |  |
| Extra small | -0.7 | -0.7 (-0.9, -0.5) | -0.1 | 0.1 (0, 0.1) | 0.9 | 1.6 (1.3, 1.8) |
| Small | -0.9 | -0.7 (-0.9, -0.5) | 0.1 | 0.2 (0.1, 0.3) | 1.2 | 1.5 (1.3, 1.7) |
| Medium | -1.2 | -1 (-1.2, -0.8) | 0.0 | 0.1 (0, 0.2) | 1.3 | 1.7 (1.4, 1.9) |
| Large | -1.3 | -1.1 (-1.4, -0.9) | 0.0 | 0.1 (0, 0.2) | 1.2 | 1.5 (1.3, 1.7) |
| Extra large | -1.5 | -1.1 (-1.5, -0.6) | -0.1 | -0.1 (-0.2, 0) | 0.9 | 0.9 (0.6, 1.2) |
| CBSA type |  |  |  |  |  |  |
| Metro | -1.2 | -1.1 (-1.3, -0.9) | 0.0 | 0.1 (0.1, 0.2) | 1.2 | 1.7 (1.5, 1.8) |
| Micro | -0.6 | -0.7 (-0.9, -0.5) | 0.0 | 0.1 (0, 0.2) | 1.0 | 1.5 (1.2, 1.8) |
| Rural | -0.7 | -0.9 (-1.1, -0.7) | -0.2 | 0 (-0.1, 0.1) | 0.7 | 1.1 (0.9, 1.4) |
| Ownership |  |  |  |  |  |  |
| For-profit | -1.0 | -0.9 (-1.1, -0.7) | 0.0 | 0.1 (0, 0.2) | 1.4 | 1.5 (1.3, 1.7) |
| Non-profit | -1.0 | -0.9 (-1.1, -0.8) | -0.1 | 0.1 (0, 0.1) | 1.1 | 1.4 (1.2, 1.5) |
| Teaching class |  |  |  |  |  |  |
| Major | -1.5 | -1 (-1.6, -0.5) | 0.0 | 0.2 (0, 0.3) | 1.7 | 2 (1.6, 2.3) |
| Minor | -1.1 | -0.8 (-0.9, -0.6) | 0.0 | 0 (0, 0.1) | 1.1 | 1.1 (1, 1.3) |
| Non-teaching | -0.9 | -0.9 (-1.1, -0.8) | -0.1 | 0 (0, 0.1) | 1.1 | 1.2 (1, 1.4) |
| Safety net status |  |  |  |  |  |  |
| Non-safety net | -1.0 | -1 (-1.2, -0.9) | -0.1 | -0.1 (-0.2, 0) | 1.0 | 1 (0.9, 1.2) |
| Safety net | -0.7 | -0.8 (-1, -0.6) | 0.2 | 0.2 (0.1, 0.3) | 1.8 | 1.8 (1.6, 2.1) |
| **Education inclusivity score** | | | | | | |
| Hospital size |  |  |  |  |  |  |
| Extra small | -0.6 | -0.9 (-1.1, -0.7) | -0.1 | 0 (-0.1, 0.1) | 1.0 | 1.3 (1.1, 1.6) |
| Small | -0.9 | -0.9 (-1.1, -0.7) | 0.0 | 0.1 (0, 0.1) | 1.3 | 1.3 (1.1, 1.6) |
| Medium | -1.1 | -1.1 (-1.3, -0.8) | 0.0 | 0 (-0.1, 0.1) | 1.3 | 1.2 (1, 1.5) |
| Large | -1.4 | -1.4 (-1.7, -1.1) | -0.1 | -0.1 (-0.2, 0) | 1.2 | 1.1 (0.9, 1.4) |
| Extra large | -1.5 | -1.2 (-1.6, -0.9) | -0.3 | -0.3 (-0.5, -0.2) | 0.8 | 0.6 (0.2, 1) |
| CBSA type |  |  |  |  |  |  |
| Metro | -1.2 | -1.3 (-1.5, -1.2) | 0.0 | 0.1 (0, 0.1) | 1.3 | 1.6 (1.4, 1.8) |
| Micro | -0.6 | -0.9 (-1.1, -0.8) | -0.1 | -0.1 (-0.2, 0) | 0.9 | 1.1 (0.9, 1.4) |
| Rural | -0.6 | -1 (-1.2, -0.9) | -0.2 | -0.2 (-0.3, -0.1) | 0.5 | 0.6 (0.4, 0.9) |
| Ownership |  |  |  |  |  |  |
| For-profit | -1.0 | -1.1 (-1.3, -0.9) | 0.0 | -0.1 (-0.2, 0) | 1.4 | 1.2 (0.9, 1.5) |
| Non-profit | -0.9 | -1.1 (-1.2, -1) | -0.1 | -0.1 (-0.1, 0) | 1.1 | 1.1 (0.9, 1.2) |
| Teaching class |  |  |  |  |  |  |
| Major | -1.9 | -1.5 (-1.9, -1.1) | -0.2 | -0.1 (-0.2, 0) | 1.1 | 1.3 (1, 1.7) |
| Minor | -1.2 | -0.9 (-1.1, -0.8) | -0.1 | -0.1 (-0.2, 0) | 1.1 | 1 (0.8, 1.2) |
| Non-teaching | -0.8 | -0.9 (-1, -0.7) | -0.1 | -0.1 (-0.1, 0) | 1.1 | 1.1 (0.9, 1.3) |
| Safety net status |  |  |  |  |  |  |
| Non-safety net | -1.0 | -1.2 (-1.3, -1) | -0.1 | -0.2 (-0.3, -0.2) | 1.0 | 0.8 (0.6, 1) |
| Safety net | -0.7 | -1 (-1.2, -0.9) | 0.1 | 0.1 (0, 0.1) | 1.8 | 1.5 (1.2, 1.7) |
| **Racial inclusivity score** | | | | | | |
| Hospital size |  |  |  |  |  |  |
| Extra small | -0.7 | -0.3 (-0.6, 0) | 0.3 | 0.3 (0.3, 0.4) | 0.4 | 1.0 (0.6, 1.3) |
| Small | -1.4 | -0.4 (-0.8, -0.1) | 0.2 | 0.3 (0.2, 0.3) | 0.5 | 1.0 (0.6, 1.3) |
| Medium | -1.4 | -0.2 (-0.5, 0.1) | 0.2 | 0.3 (0.3, 0.4) | 0.6 | 1.0 (0.6, 1.4) |
| Large | -1.2 | -0.1 (-0.5, 0.3) | 0.2 | 0.4 (0.4, 0.4) | 0.9 | 1.1 (0.7, 1.5) |
| Extra large | -1.2 | 0 (-0.5, 0.5) | 0.3 | 0.4 (0.4, 0.5) | 0.7 | 0.9 (0.6, 1.3) |
| CBSA type |  |  |  |  |  |  |
| Metro | -1.5 | -1 (-1.2, -0.8) | 0.2 | 0.2 (0.2, 0.3) | 0.6 | 1.0 (0.6, 1.4) |
| Micro | -0.5 | 0.1 (-0.2, 0.4) | 0.3 | 0.4 (0.4, 0.4) | 0.4 | 1.0 (0.6, 1.4) |
| Rural | -0.2 | 0.2 (-0.1, 0.5) | 0.3 | 0.4 (0.4, 0.4) | 0.4 | 1.0 (0.6, 1.4) |
| Ownership |  |  |  |  |  |  |
| For-profit | -0.8 | -0.1 (-0.3, 0.2) | 0.3 | 0.4 (0.3, 0.4) | 0.7 | 1.0 (0.7, 1.4) |
| Non-profit | -1.2 | -0.4 (-0.6, -0.1) | 0.2 | 0.3 (0.3, 0.3) | 0.5 | 1.0 (0.6, 1.3) |
| Teaching class |  |  |  |  |  |  |
| Major | -0.8 | 0.1 (-0.5, 0.7) | 0.3 | 0.4 (0.4, 0.5) | 1.5 | 1.4 (0.3, 2.4) |
| Minor | -1.2 | -0.3 (-0.5, -0.1) | 0.2 | 0.3 (0.3, 0.4) | 0.6 | 0.8 (0.7, 1) |
| Non-teaching | -1.1 | -0.5 (-0.7, -0.3) | 0.2 | 0.3 (0.3, 0.3) | 0.4 | 0.8 (0.7, 0.9) |
| Safety net status |  |  |  |  |  |  |
| Non-safety net | -1.3 | -0.5 (-0.7, -0.3) | 0.2 | 0.3 (0.3, 0.3) | 0.4 | 0.6 (0.3, 1) |
| Safety net | -0.3 | 0.1 (-0.2, 0.3) | 0.3 | 0.4 (0.4, 0.4) | 1.3 | 1.4 (1, 1.8) |

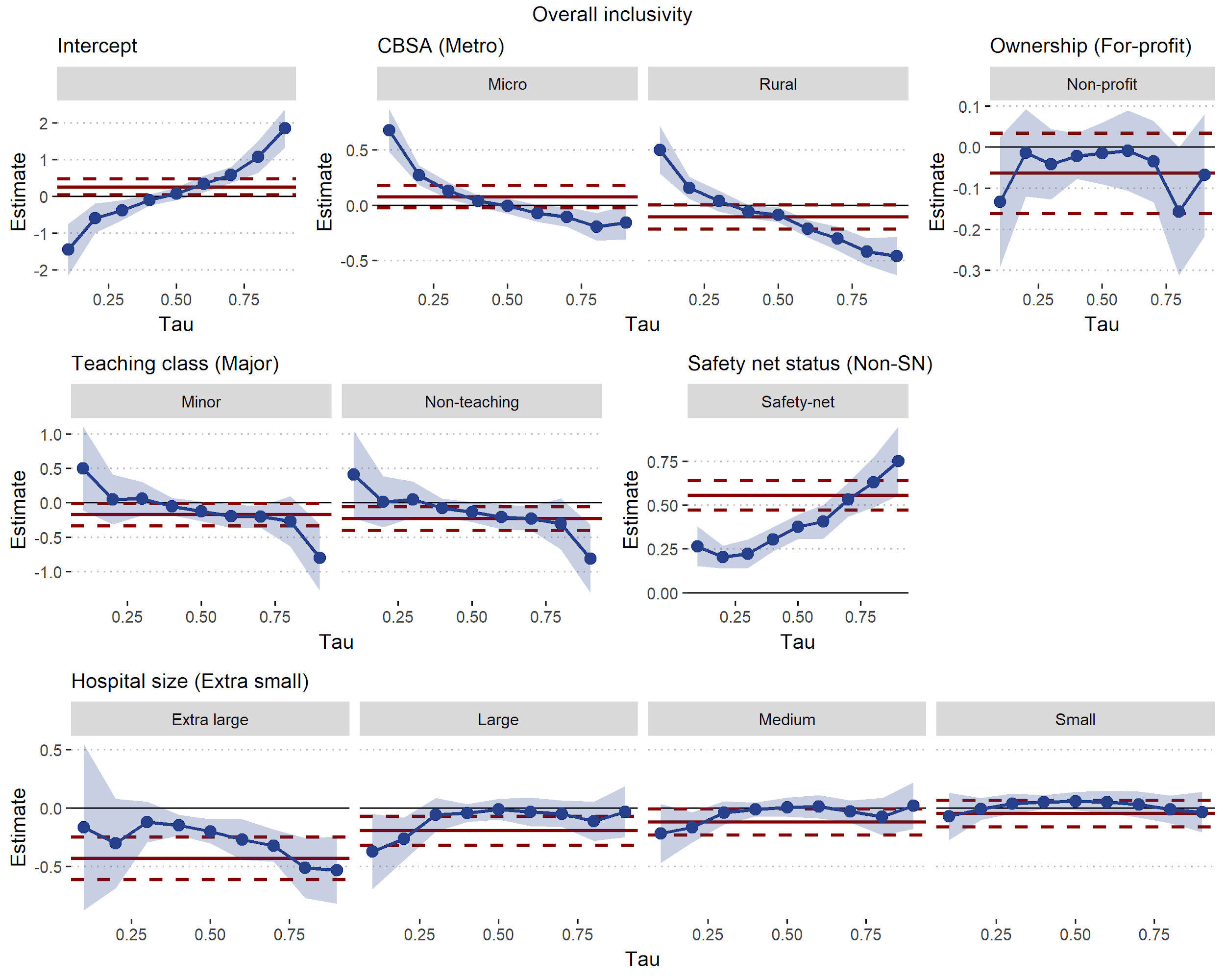

### Appendix Figure 1. The hospital characteristic estimates and 95% confidence intervals (CI) from quantile regressions (blue lines, shaded area is 95% CI) and from an ordinary least squares regression (solid red lines, dashed lines is 95% CI) for the overall inclusivity score. The comparator hospital characteristic is reported in brackets. Tau represents the predicted quantile from each quantile regression.

A.

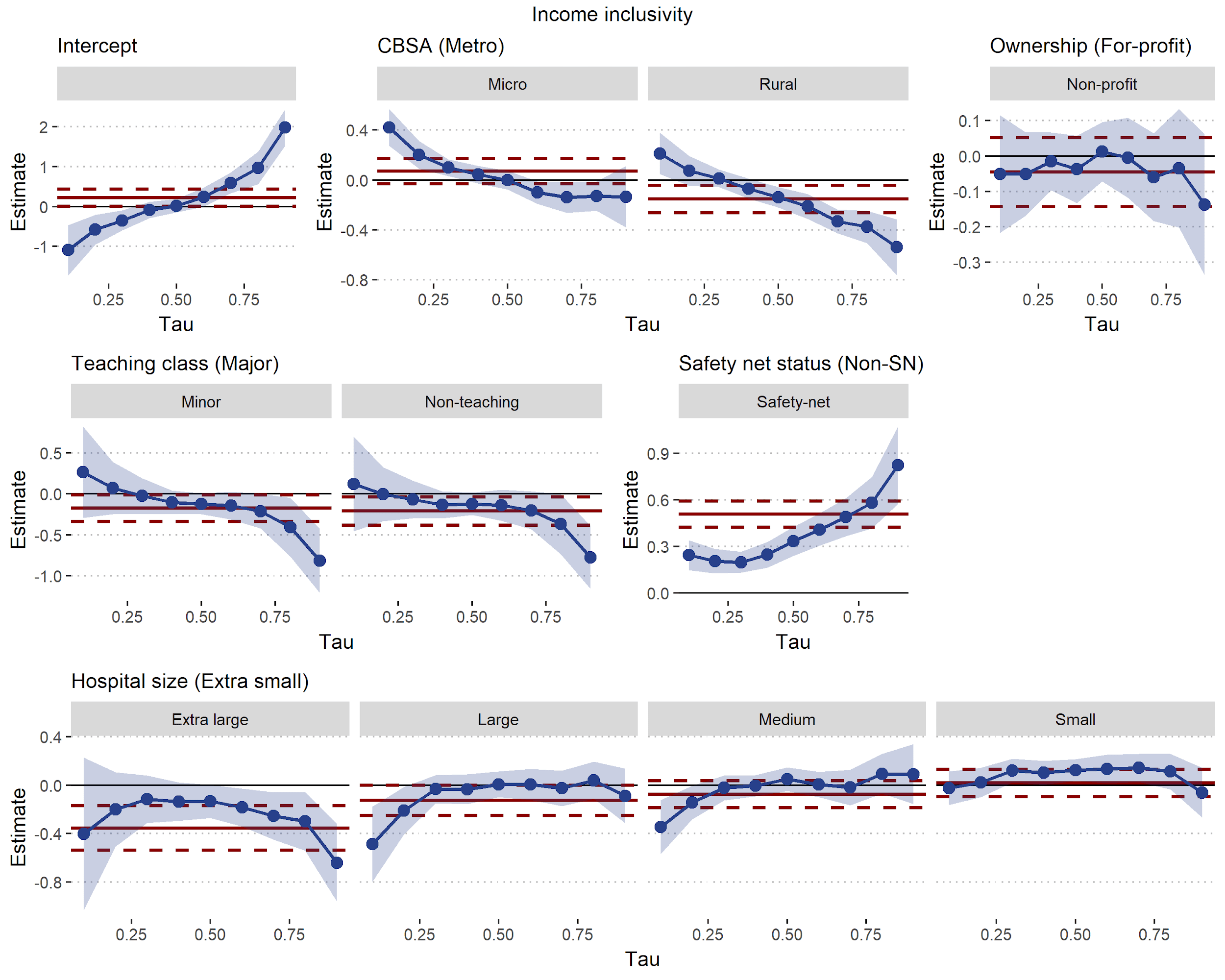

B.

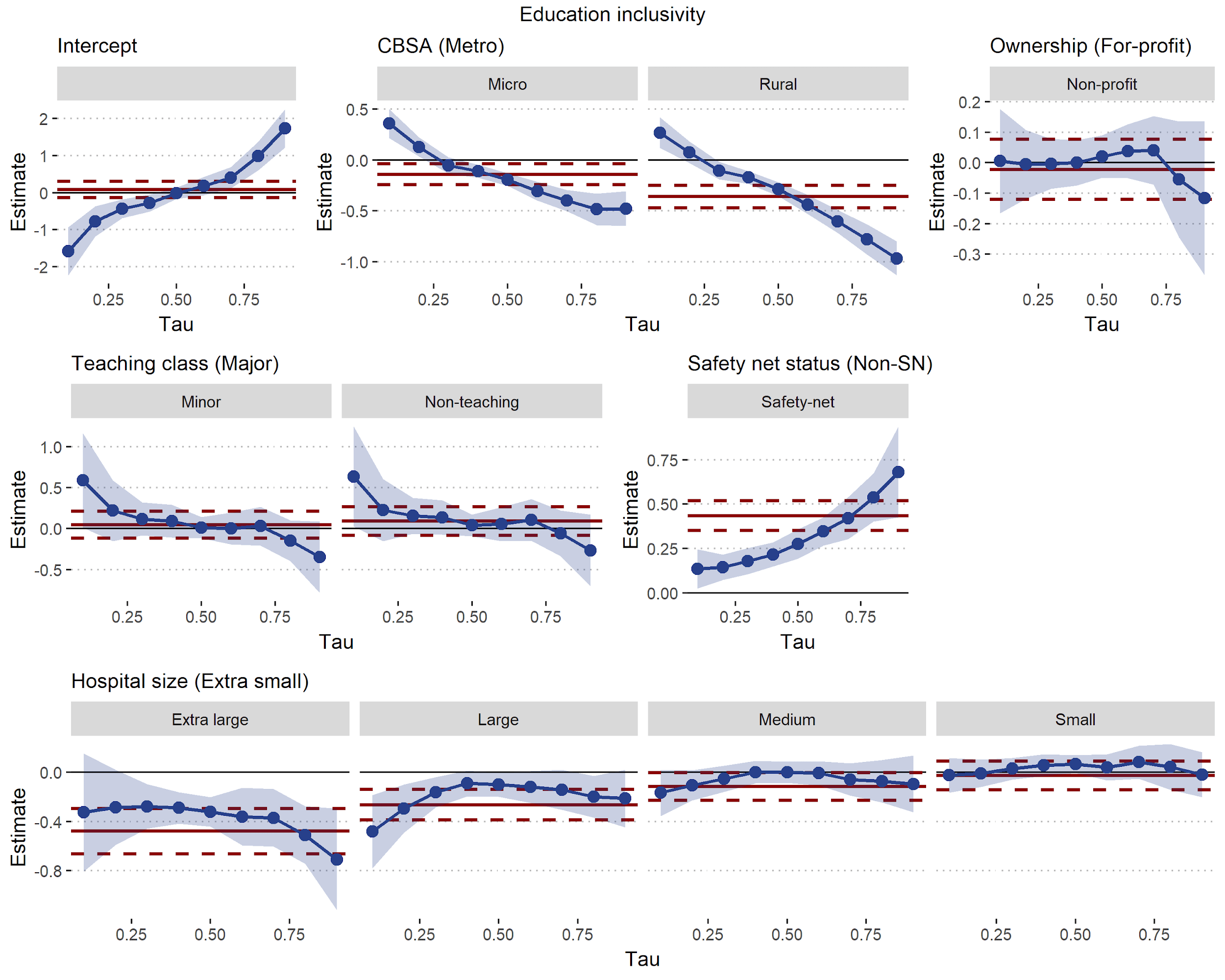

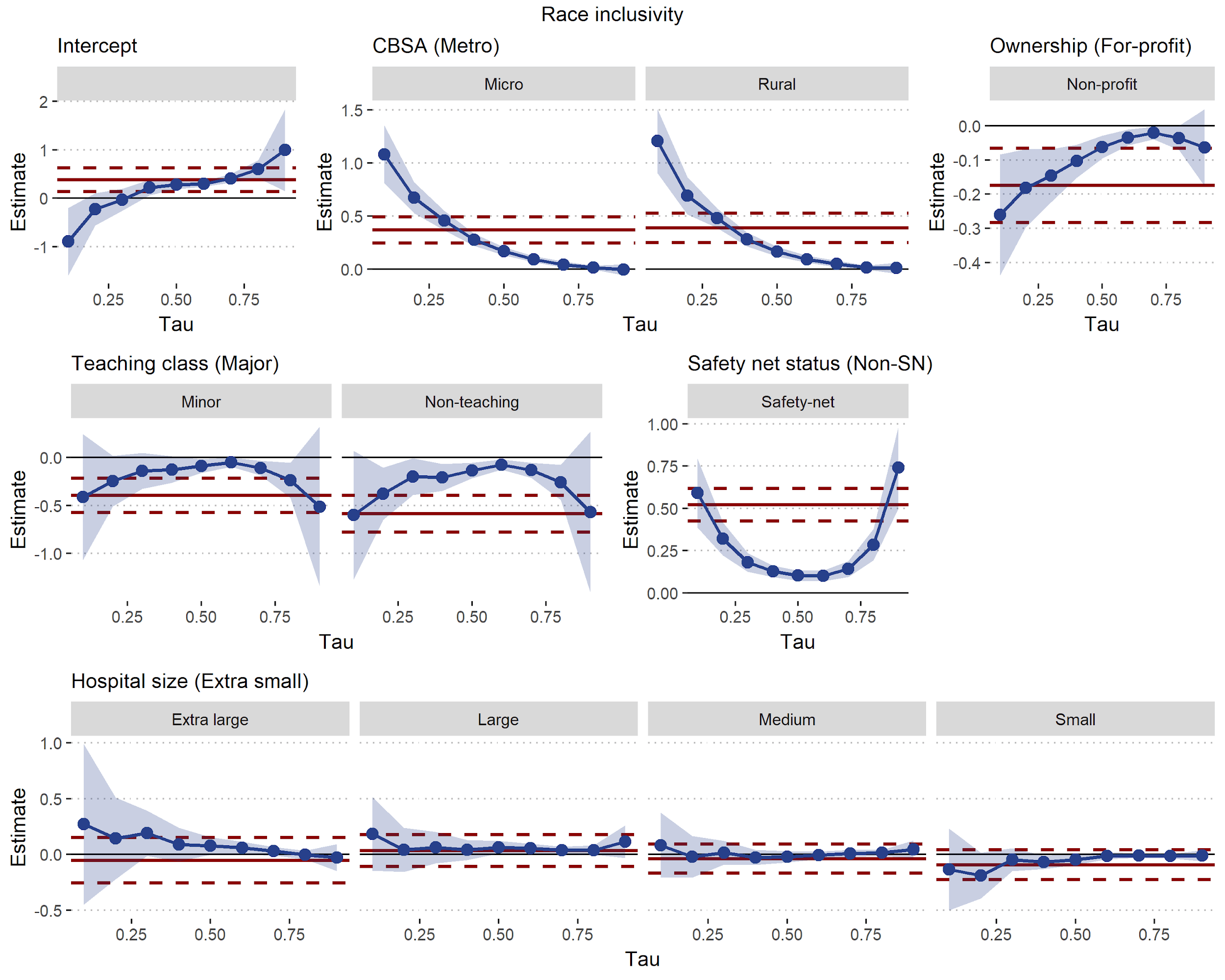

C.

### Appendix Figure 2. Quantile regression results of inclusivity scores for income (A), education (B) and racial inclusivity (C).

### Appendix Table 4. Comparison of inclusivity scores of safety net versus non safety net hospitals in rural areas.

| **Characteristic** | **Non safety net hospitals, N = 518** | **Safety net hospitals, N = 201** | **p-value^1^** |
| --- | --- | --- | --- |
| Overall inclusivity |  |  | 0.002 |
| Median (IQR) | -0.1 (-0.3, 0.2) | 0.0 (-0.2, 0.5) |  |
| Mean (SD) | 0.0 (0.7) | 0.1 (0.6) |  |
| Racial inclusivity |  |  | 0.057 |
| Median (IQR) | 0.3 (0.1, 0.3) | 0.3 (0.2, 0.3) |  |
| Mean (SD) | 0.2 (0.7) | 0.3 (0.3) |  |
| Income inclusivity |  |  | 0.014 |
| Median (IQR) | -0.2 (-0.4, 0.2) | -0.1 (-0.3, 0.4) |  |
| Mean (SD) | -0.1 (0.7) | 0.1 (0.7) |  |
| Education inclusivity |  |  | 0.008 |
| Median (IQR) | -0.2 (-0.4, 0.1) | -0.1 (-0.3, 0.2) |  |
| Mean (SD) | -0.1 (0.6) | 0.0 (0.6) |  |
| ^1^Welch Two Sample t-test | | | |

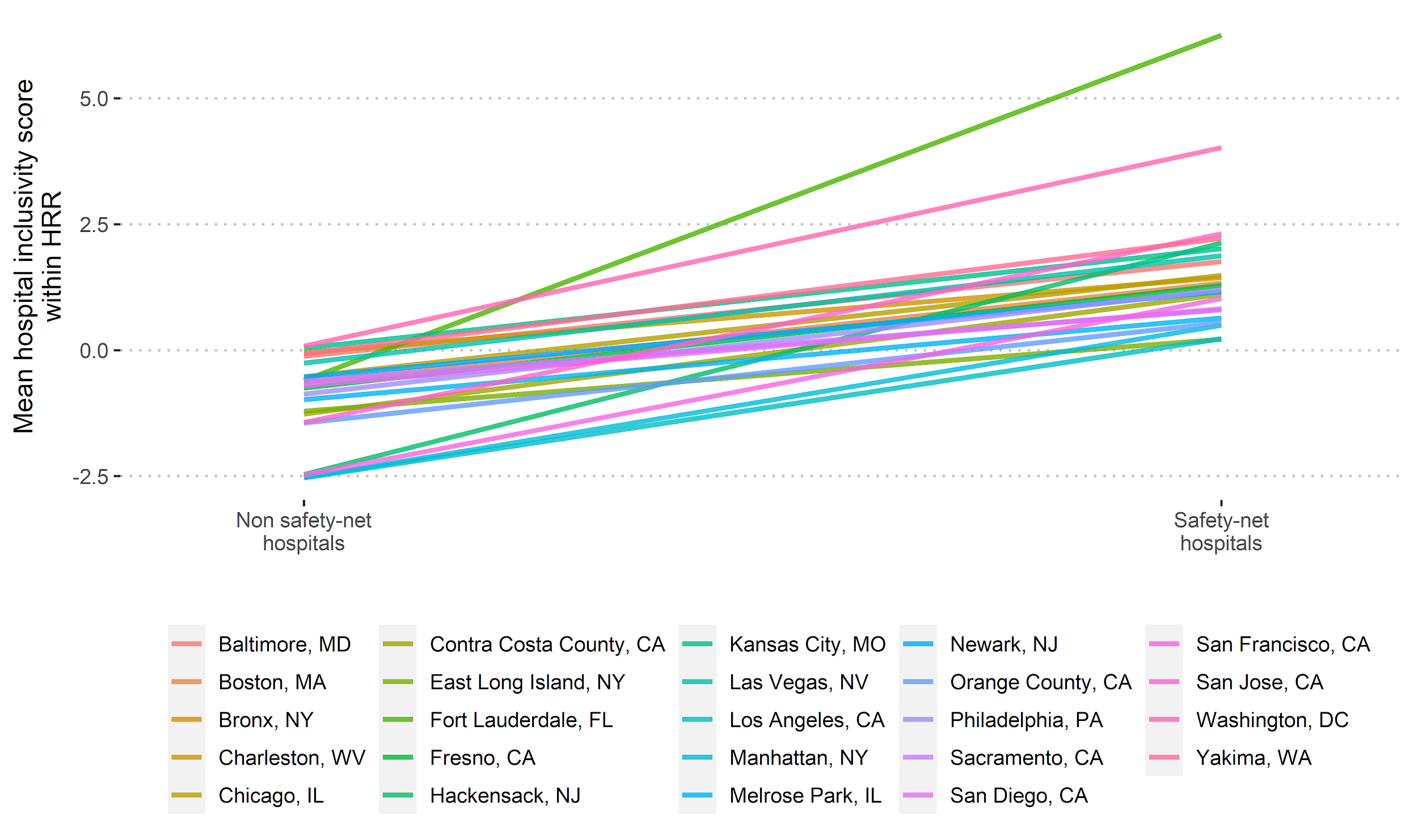

### Appendix Figure 3. The mean overall inclusivity scores for safety net hospitals and non-safety net hospitals for selected hospital referral regions (HRR). These are the HRRs where there was a significant slope coefficient for safety net status in a multi-level regression with HRR as a random effect.

### **Additional Inclusivity Methods**

***Specialty hospital definition***

Specialty hospitals were defined as those with more than 40% of their annual admissions as either orthopedic Diagnosis Related Groups (DRG), cardiac DRGs, or admissions with a surgical International Classification of Disease 10^th^ edition code and an elective procedure flag.

***Selecting furthest zip code tabulation area from hospital***

For each campus location, we sorted ZCTAs by descending patient counts and then calculated the cumulative percentage of patients by these ordered ZCTAs. Conceptually, we wanted to find the ordered ZCTA along this curve (the cumulative percentage of patients) at which the addition of more patients made a small difference to the total patient count. We found this ‘turning point’ as the point along this ZCTA versus cumulative percentage curve that was furthest from a straight line drawn from the start to end points of the curve. We then selected the ZCTA at this turning point (referred to from here as ZCTA *A*).

We then calculated the geodesic distance from the hospital address to the centroid of all ZCTAs where the hospital had patients.^1^ We examined all ZCTAs which had an equal or greater patient count compared to ZCTA *A*, and selected the maximum distance from the hospital. To account for noise and small patient counts from some ZCTAs, we made one adjustment: we found the minimum distance across all ZCTAs that had patient counts within ±1 of ZCTA *A*’s patient count. This distance alone was compared to the distances of ZCTAs where the patient count was greater than ZCTA *A*’s patient count, plus one. This prevented a distant ZCTA with a small count of patients from being selected as the final distance. Once we found this CA-R, we found the distance between each hospital address and all ZCTA centroids within this radius, including ZCTAs with zero patient counts.

### **Results of Quantile Regression for Each Inclusivity Measure**

*Income Inclusivity*

At the 90^th^ percentile of income inclusivity scores (Appendix Figure 2A) major teaching versus non-teaching was 0.61 points greater (adjusted 0.82, CI: 0.39 to 1.24; p = 0.001) and 0.58 points greater (adjusted 0.78 CI: 0.34 to 1.21; p < 0.001) versus minor teaching hospitals. The 10^th^ percentile score difference between major teaching hospitals was -0.63 points for non-teaching (adjusted -0.12, CI: -0.80 to 0.56; p = 0.91) and -0.36 points for minor-teaching (adjusted -0.27, CI: -0.94 to 0.41; p = 0.63) hospitals. Extra-large hospitals had the lowest 90^th^ percentile income inclusivity scores (adjusted value 0.93, CI: 0.65 to 1.22), compared to other hospital sizes.

*Education Inclusivity*

For education inclusivity (Appendix Figure 3B), the hospitals with the highest 90th percentile scores were metro-area, safety net and extra-large hospitals. The hospitals with the lowest 10^th^ percentile education inclusivity scores were metro-area, non-safety net hospitals and major teaching hospitals. The difference in the 10^th^ percentile education inclusivity score for major teaching hospitals versus minor was -0.75 points (adjusted -0.64, CI: -1.13, -0.05 points; p = 0.029) and was -1.13 points (-0.64, CI: -1.18 to -0.09; p = 0.018) for non-teaching hospitals.

*Racial Inclusivity*

The highest 90^th^ percentile scores for racial inclusivity were for safety net hospitals (compared to non-safety net; Appendix Figure 3A). Unlike the income and education scores, at the 90^th^ percentile scores for race, there were no differences between metro micro or rural areas.

The hospitals with the lowest 10^th^ percentile racial inclusivity scores were metro-area hospitals, nonprofit hospitals and non-safety net hospitals. The difference in the 10th percentile score for metro- versus rural area hospitals was -1.25 points (adjusted -1.21, CI: -1.57 to -0.85; p < 0.001), and versus micro-area hospitals was -1.01 points (adjusted -1.08, CI: -1.42 to -0.75; p < 0.001).

The only difference between nonprofit and for-profit hospitals was found at the 10^th^ percentile (lowest scores) of racial inclusivity, where the for-profit hospital score was 0.35 points greater than non-profit hospitals (adjusted difference of 0.26, CI: 0.08 to 0.44, p = 0.004).

### **Additional Discussion Points**

Gresenz and colleagues^2^ found that rural hospital market radii were larger than urban hospitals. We found the opposite when comparing metropolitan hospitals to rural hospitals, using our travel time radius and the distance of the furthest ZCTA in the community area. This difference may be due to our use of CBSA definitions of population areas rather than quintiles of population density. We did note that our radius was dependent on hospital size, and as most rural hospitals were small or extra small hospitals their radius was small. Micropolitan areas had the greatest median CA travel radius, as well as a greater proportion of their total patients within the CA, suggesting that hospitals in these intermediate population zones may serve a particular function as magnets for surrounding areas.
